# Cardiometabolic risk factors and preclinical target organ damage among adults in Ghana: Findings from a national study

**DOI:** 10.1101/2020.04.28.20082446

**Authors:** Jie Li, Isaac Kofi Owusu, Qingshan Geng, Aba Ankomaba Folson, Zhichao Zheng, Yaw Adu-Boakye, Xinran Dong, Wen-Chih Wu, Francis Agyekum, Hongwen Fei, Harold Ayetey, Mulan Deng, Fred Adomako-Boateng, Zuxun Jiang, Braimah Baba Abubakari, Zhao Xian, Forster Nketiah Fokuoh, Lambert Tetteh Appiah, Simin Liu, Chunying Lin

## Abstract

**Background:** Sub-Saharan Africa (SSA) has the highest prevalence of cardiovascular diseases (CVD). Nevertheless, very few studies have directly examined the development of and risk factors for CVD among Africans.

**Objective:** To examine CVD risk factors and outcomes particularly in the early stage of CVD development among adults in Ghana.

**Methods:** Using a stratified multistage random sampling method, 1,106 participants were recruited as a nationally representative sample of the general population ≥18 years in Ghana from 2016 to 2017. For each participant, we measured CVD risk factors and preclinical target organ damage (TOD) for CVD development.

**Results:** The prevalence of CVD risk factors was 21.1% for obesity, 10.8% for diabetes, 55.4% for hypertension, 37.3% for dyslipidemia, 12.8% for hyperuricemia, and 39.3% for hsCRP>3 mg/L in the recruited population. The prevalence of preclinical TOD was 8.6% for peripheral artery disease (PAD), 14.7% for carotid thickening, 5.9% for left ventricular hypertrophy (LVH), and 4.4% for chronic kidney disease (CKD). Three CVD risk factors appeared to play most prominent roles in TOD development, including obesity for PAD (OR 1.88, 95% CI 1.13–3.09), hypertension for carotid thickening (OR 1.57, 95% CI 0.99–2.54) and LVH (OR 6.25, 95% CI 2.98–14.50), and hyperuricemia for CKD (OR 5.56, 95% CI 2.79–11.15).

**Conclusions:** The prevalence of CVD risk factors and early outcomes have reached epidemic proportions among Ghanaian adults. The distinct patterns of risk factors in the development of TOD presents important challenges and opportunities for interventions to improve cardiometabolic health among adults in Ghana.

## Introduction

Globally, cardiovascular diseases (CVD) remain the leading cause of death. However, the CVD burden has not changed uniformly across world regions of different socioeconomic characteristics. Over the past decades, CVD mortality has dramatically declined in high-income countries, while increased in low- and middle- income countries (LMICs)^1^. According to the Global Burden Disease (GBD) study in 2015, the age-standardized CVD prevalence is 6, 304 per 100,000 globally with the highest rate in western sub-Saharan Africa (SSA) (9,475 per 100,000) ^2^. There is a paucity of data on CVD epidemiology, prevention, and treatment in many LMICs, especially in SSA^3^. Thus, our understanding on CVD is disproportionately informed by studies conducted in high-income countries, findings of which may not be directly applicable to peoples in LMICs. Given that the genetic characteristics and CVD risk factors differ across ethnicities and regions, systemic and comprehensive studies on CVD and the relevant risk factors in SSA is urgently needed to guide proper allocation of health care resource and public health policy for the region.

CVD is a multistage pathogenetic disorder spanning lifetimes, often affected by a cluster of risk factors including obesity, dyslipidemia, diabetes, and hypertension. Preclinical pathological changes in blood vessels, the heart and kidneys, termed target organ damage (TOD), are common and typically lead to end-stage organ failure in the absence of intervention^4^. Although often asymptomatic, preclinical TOD can be considered intermediate endpoints for CVD events in the pathogenesis of CVD. There is now a body of accumulating evidence that indicates CVD progression can be modified by earlier interventions^5^.

In the current study, we conducted a population-based study in Ghana located in SSA aimed to examine the prevalence of CVD risk factors and preclinical TOD using a national representative sample of the general population. Specifically, we investigated whether and to what extent the known CVD risk factors can lead to preclinical vascular, cardiac, and renal TOD among adults in Ghana.

## Methods

### Study participants

In this Ghana Heart Study, we used a stratified three-stage random sampling strategy to recruit a nationally representative sample of the general Ghanaian population 18 years of age or older. Stratification was based on geographic region with representation of the southern (Greater Accra and Central regions), middle (Ashanti regions) and northern (Northern regions) zones of the country. Each of the four regions was divided into urban areas and rural areas on the basis of administrative data. On the first stage of selection, one urban and one rural community were selected by simple random sampling method from the urban and rural areas in each region, respectively. The second stage of sample selection consisted of households. In a given community, a listing of all households was prepared, and a subsample of these was selected by a systematic sampling method. The third stage of sample selection consisted of persons within the selected households. All members ≥ 18 years within a household were listed, and a subsample of individuals was selected by simple random sampling method based on sex, age, ethnicity, and income. We excluded participants who were pregnant, had type 1 diabetes mellitus, or refused to give consent. A total of 1,106 participants completed the study and were included in the final analysis.

All procedures were carried out according to the study protocol approved by the Committee on Human Research Publication and Ethics of School of Medicine and Dentistry, the Kwame Nkrumah University of Science and Technology, Kumasi, Ghana, and Research Ethics Committee of Guangdong Provincial People’s Hospital, Guangdong Academy of Medical Sciences, Guangzhou, China. Informed consent was obtained from all the participants. The objectives and nature of the study were explained to all participants. This study was registered at www.chictr.org.cn as ChiCTR1800017374.

### Cardiometabolic risk factors measurements

A standard questionnaire was used to obtain information on demographics, personal and family medical history, and lifestyle risk factors including smoking, drinking, and physical activity. Adequate physical activity was defined as at least 150 minutes per week of moderate or vigorous activities such as brisk walking.

Body weight, height, and waist circumference were measured with light clothes and bare feet. Body mass index (BMI) was calculated as weight in kilogram divided by the square of the height in meters. Blood pressure was measured using the OMRON M6 devices with appropriate cuff sizes. Three blood pressure readings were taken from the left arm, with participants in the sitting position after a 10-minutes of inactivity. The mean of the recorded readings was taken as the participant’s blood pressure. Fasting plasma samples were collected to measure glucose, HbA1c, total triglyceride (TG), total cholesterol (TC), low density lipoprotein (LDL) cholesterol, high density lipoprotein (HDL) cholesterol, lipoprotein (a)[Lp(a)], hsCRP, creatinine (Cr), and uric acid. The criteria used for diagnosing obesity, diabetes, hypertension, dyslipidemia, high hsCRP, and hyperuricemia were provided in supplemental methods.

### Preclinical TOD measurements

Each participant underwent physical examinations to evaluate for preclinical vascular, cardiac, and renal TOD. For vascular TOD, ankle brachial pressure index (ABI) was measured for each participant using the Boso (Bosch & Sohn, Germany) ABI device. Carotid ultrasound was performed for both carotid arteries using the GE VIVID Q ultrasound system equipped with a 9L-RS 3.1-10.0 MHz linear probe to determine carotid intima media thickness (CIMT). For cardiac TOD, each participant had a transthoracic echocardiography examination by a cardiologist using the GE VIVID Q ultrasound system equipped with M4S-RS 1.5-3.5 MHz sector probe. Left ventricular mass (LVM) was corrected for body surface area (BSA), which was termed left ventricular mass index (LVMI). Renal TOD was evaluated using the estimated glomerular filtration rate (eGFR), an important chronic kidney disease (CKD) marker. The criteria used for diagnosing peripheral artery disease (PAD), Carotid intimal thickening, left ventricular hypertrophy (LVH), and CKD were provided in supplemental methods.

### Statistical analysis

Basic characteristics, cardiometabolic risk factors, and preclinical TOD measurements were described by sex and ethnicity. Continuous variables were presented as means and standard deviations (SD) for those with normal distribution or medians and interquartile ranges (IQR) for those with non-normal distribution. Categorical variables were shown in percentages.

To increase the statistical power, we treated cardiometabolic risk factors and TOD measurements as continuous and employed multivariable linear regression model to analyze the association between risk factors and TOD measurements. For the analysis of BMI and waist circumference with TOD measurements, age, sex, ethnicity, living area, education, smoking, and physical activity were adjusted in the model. For the other risk factors, BMI was additionally controlled in the model. Coefficients and 95% CIs (confidence intervals) were calculated in the linear regression model. Stratification analyses by sex and ethnicity were conducted to evaluate sex and ethnicity differences for the risk factor-TOD association.

The association between categorized risk factors and preclinical TOD was analyzed with the logistic regression model. The model for the association of overall obesity and abdominal obesity with TOD, adjusted for age, sex, ethnicity, living area, education, smoking, and physical activity. For the analysis of other risk factors, BMI was further controlled in the model. The associations of risk factors with TOD were presented as odd ratios (ORs) and 95% CIs.

Missing values for covariates were imputed using multivariate imputation by chained equations^6^.A 2-sided *P* < 0.05 was considered statistically significant. All analyses were conducted with the use of R 3.5.2 software (The R Foundation for Statistical Computing).

## Results

### Basic characteristics of study participants in Ghana

As shown in Table 1, of the 1,106 participants aged 46.9 ± 17.2 years old,42% were men, and 58% were Akan. Compared with women, more men were educated higher than middle school (70% vs 47%), smokers (7.8% vs 1.2%), to consume alcohol (57% vs 44%), and be physically active (22% vs 12%). Compared with the other ethnicities, Akan were more likely to be educated higher than middle school (60% vs 53%), to consume alcohol (62% vs 31%), and were less physically active (9% vs 26%).

**Table 1.**
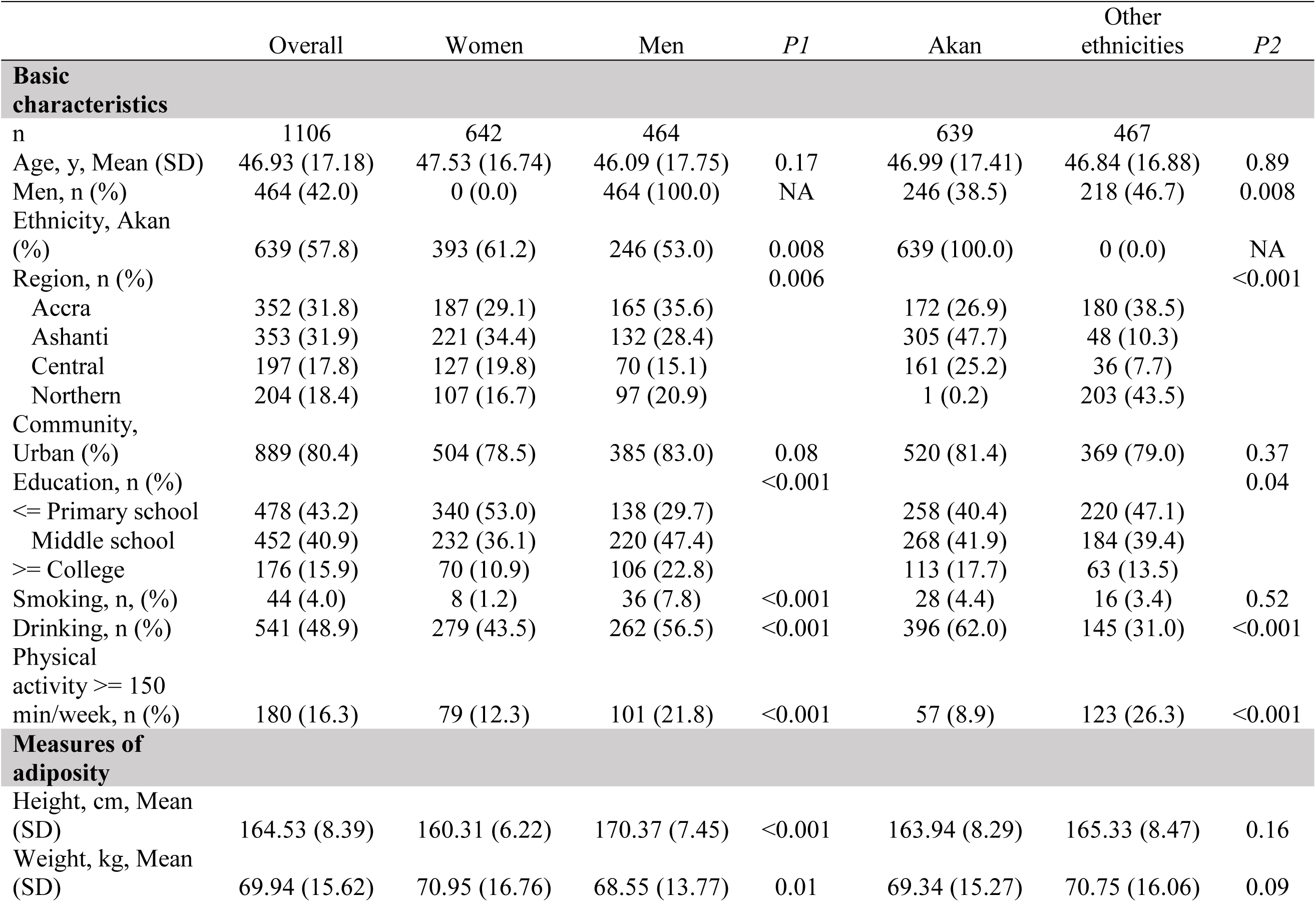
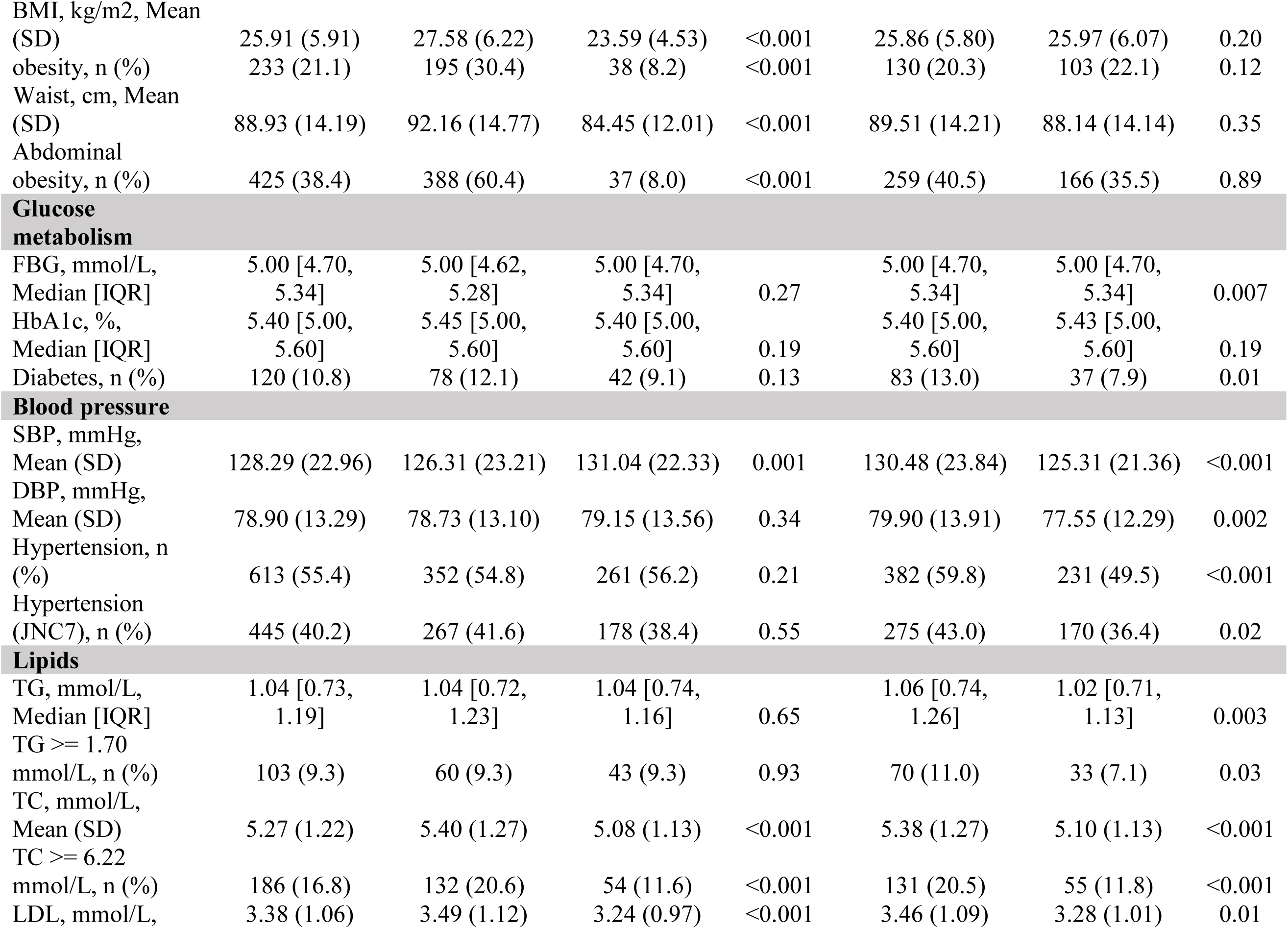
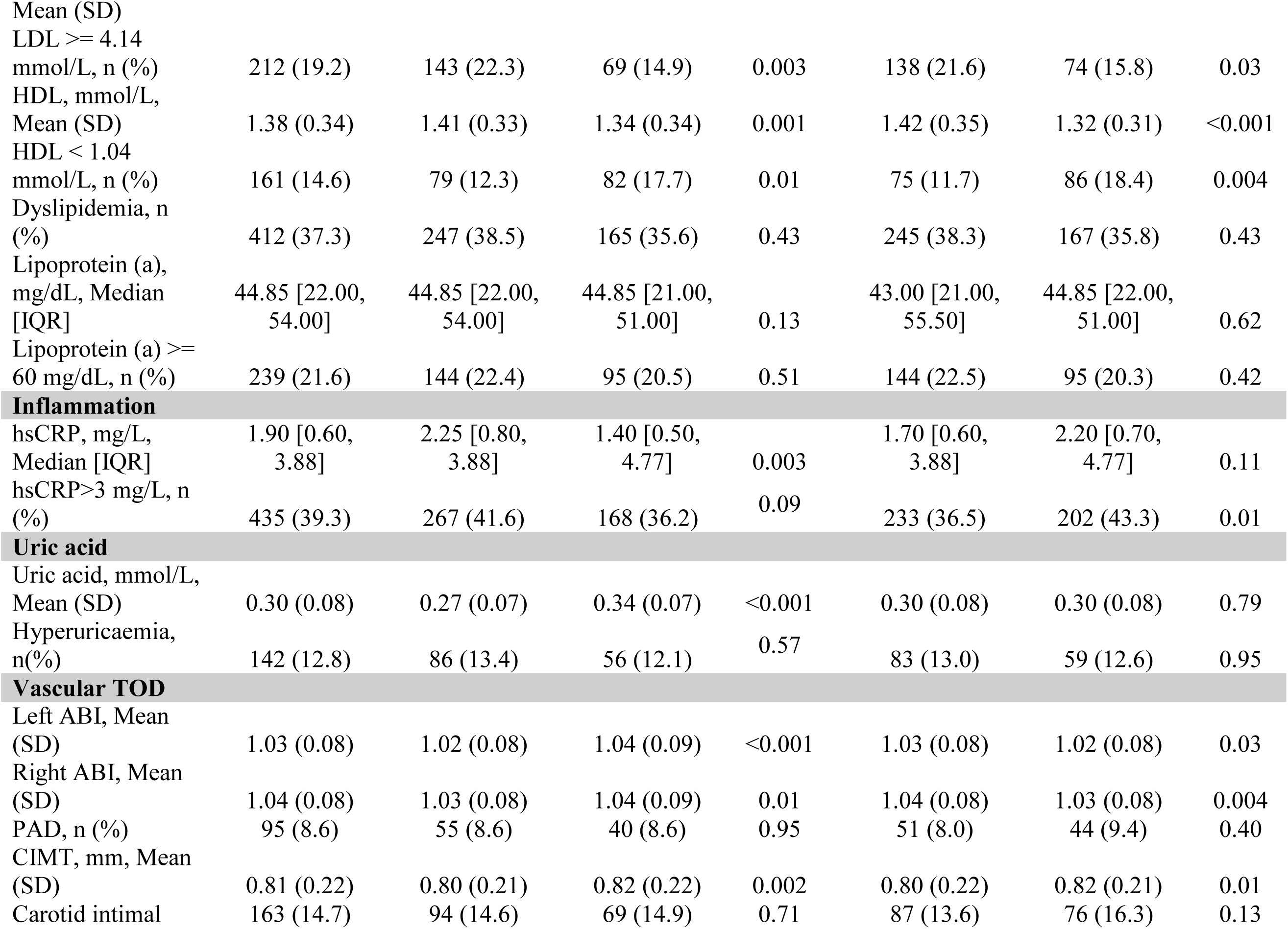
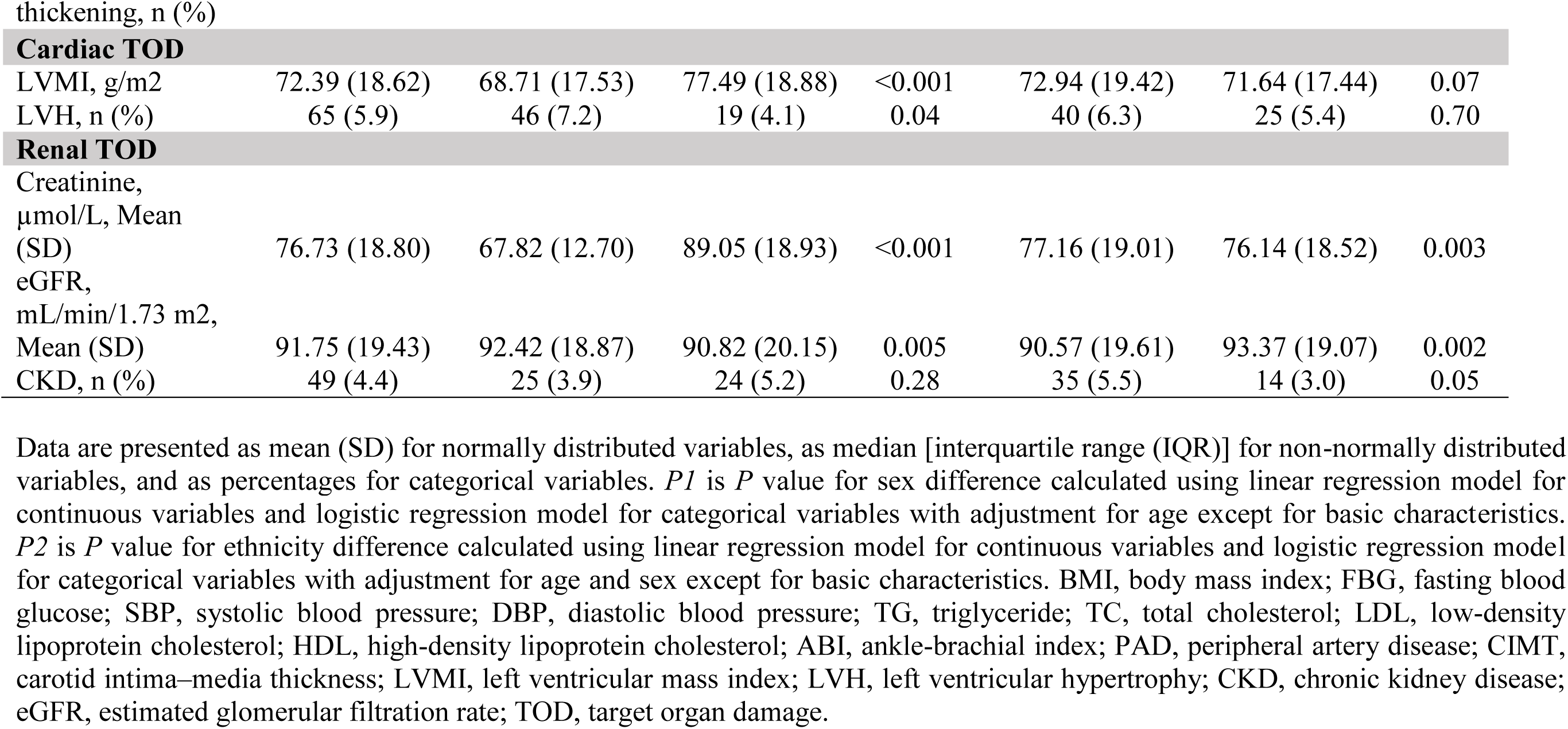
Cardiometabolic characteristics of adults participating in the Ghana Heart Study, 2016–2017

### Cardiometabolic risk factors of study participants in Ghana

In this nationally representative population, 21% were overall obese, 38% had abdominal obesity, 55% were hypertensive, 11% had diabetes, 37% had dyslipidemia, 13% had hyperuricemia, and 39% had an hsCRP higher than 3 mg/L. Compared with men, women were significantly more obese (overall obesity, 30% vs 8%, *P* < 0.001; abdominal obesity, 60% vs 8%, *P* < 0.001), had higher plasma TC, LDL, HDL, and hsCRP levels, but lower systolic blood pressure (SBP)and uric acid levels. Compared with the other ethnicities, Akan had higher fasting blood glucose, blood pressure, and lipids, but less hsCRP (Table 1).

### Preclinical TOD of study participants in Ghana

As shown in Table 1, 8.6% had PAD, 15% had carotid thickening, 5.9% had LVH, and 4.4% had CKD. Women had significantly higher LVH than men (7% vs 4%, *P*=0.04). Akan had higher CKD than the other ethnicities (5.5% vs 3%, *P*=0.05).

### The association of cardiometabolic risk factors with preclinical TOD

Among these known risk factors, obesity had the strongest association with PAD (OR 1.88, 95%CI 1.13–3.09), independent from age, sex, ethnicity, region, living community, education, smoking, and physical activity(Table 2). Stratification analysis showed that the association between BMI and ABI was not affected by sex and ethnicity (Figure 1).

**Table 2.** Association between cardiometabolic risk factors and specific target organ damage (TOD) among adults participating in Ghana Heart Study, 2016–2017

|  |  | n | PAD |  |  | Carotid Intimal Thickening |  |  |
| --- | --- | --- | --- | --- | --- | --- | --- | --- |
|  |  |  | case (%) | OR (95% CI) | *P* | case (%) | OR (95% CI) | *P* |
| Overall obesity | No | 873 | 64 (7.3) | Ref |  | 125 (14.3) | Ref |  |
|  | Yes | 233 | 31 (13.3) | 1.88 (1.13, 3.09) | 0.01 | 38 (16.3) | 1.34 (0.84, 2.11) | 0.22 |
| Abdominal obesity | No | 681 | 49 (7.2) | Ref |  | 97 (14.2) | Ref |  |
|  | Yes | 425 | 46 (10.8) | 1.85 (1.08, 3.25) | 0.03 | 66 (15.5) | 0.99 (0.63, 1.58) | 0.98 |
| Diabetes | No | 986 | 83 (8.4) | Ref |  | 132 (13.4) | Ref |  |
|  | Yes | 120 | 12 (10.0) | 0.88 (0.42, 1.71) | 0.72 | 31 (25.8) | 1.30 (0.76, 2.20) | 0.33 |
| Hypertension | No | 493 | 41 (8.3) | Ref |  | 38 (7.7) | Ref |  |
|  | Yes | 613 | 54 (8.8) | 0.83 (0.50, 1.37) | 0.46 | 125 (20.4) | 1.57 (0.99, 2.54) | 0.06 |
| High TG | No | 1003 | 82 (8.2) | Ref |  | 141 (14.1) | Ref |  |
|  | Yes | 103 | 13 (12.6) | 1.32 (0.66, 2.49) | 0.41 | 22 (21.4) | 1.31 (0.71, 2.34) | 0.37 |
| High TC | No | 920 | 71 (7.7) | Ref |  | 124 (13.5) | Ref |  |
|  | Yes | 186 | 24 (12.9) | 1.50 (0.87, 2.52) | 0.13 | 39 (21.0) | 1.51 (0.93, 2.43) | 0.09 |
| High LDL | No | 894 | 65 (7.3) | Ref |  | 125 (14.0) | Ref |  |
|  | Yes | 212 | 30 (14.2) | 1.81 (1.10, 2.96) | 0.02 | 38 (17.9) | 1.20 (0.75, 1.90) | 0.45 |
| Low HDL | No | 945 | 82 (8.7) | Ref |  | 134 (14.2) | Ref |  |
|  | Yes | 161 | 13 (8.1) | 0.87 (0.44, 1.60) | 0.67 | 29 (18.0) | 1.07 (0.62, 1.83) | 0.80 |
| Dyslipidemia | No | 694 | 51 (7.3) | Ref |  | 94 (13.5) | Ref |  |
|  | Yes | 412 | 44 (10.7) | 1.23 (0.78, 1.91) | 0.37 | 69 (16.7) | 0.97 (0.65, 1.44) | 0.88 |
| High Lp(a) | No | 867 | 66 (7.6) | Ref |  | 123 (14.2) | Ref |  |
|  | Yes | 239 | 29 (12.1) | 1.40 (0.85, 2.24) | 0.17 | 40 (16.7) | 1.00 (0.63, 1.56) | 0.99 |
| High hsCRP | No | 671 | 50 (7.5) | Ref |  | 92 (13.7) | Ref |  |
|  | Yes | 435 | 45 (10.3) | 1.52 (0.97, 2.39) | 0.07 | 71 (16.3) | 0.80 (0.54, 1.18) | 0.26 |
| Hyperuricemia | No | 964 | 77 (8.0) | Ref |  | 138 (14.3) | Ref |  |
|  | Yes | 142 | 18 (12.7) | 1.13 (0.61, 1.99) | 0.70 | 25 (17.6) | 0.88 (0.50, 1.50) | 0.65 |

**Table 2 (continued)**
|  |  | n | LVH |  |  | CKD |  |  |
| --- | --- | --- | --- | --- | --- | --- | --- | --- |
|  |  |  | case (%) | OR (95% CI) | P | case (%) | OR (95% CI) | P |
| Overall obesity | No | 873 | 53 (6.1) | Ref |  | 39 (4.5) | Ref |  |
|  | Yes | 233 | 12 (5.2) | 0.79 (0.38, 1.53) | 0.50 | 10 (4.3) | 1.09 (0.48,2.31) | 0.83 |
| Abdominal obesity | No | 681 | 38 (5.6) | Ref |  | 28 (4.1) | Ref |  |
|  | Yes | 425 | 27 (6.4) | 0.83 (0.45, 1.51) | 0.54 | 21 (4.9) | 1.29 (0.61,2.75) | 0.51 |
| Diabetes | No | 986 | 57 (5.8) | Ref |  | 36 (3.7) | Ref |  |
|  | Yes | 120 | 8 (6.7) | 0.97 (0.40, 2.11) | 0.94 | 13 (10.8) | 1.38 (0.63,2.88) | 0.41 |
| Hypertension | No | 493 | 9 (1.8) | Ref |  | 5 (1.0) | Ref |  |
|  | Yes | 613 | 56 (9.1) | 6.25 (2.98, 14.5) | <0.001 | 44 (7.2) | 3.04 (1.21,9.32) | 0.03 |
| High TG | No | 1003 | 63 (6.3) | Ref |  | 41 (4.1) | Ref |  |
|  | Yes | 103 | 2 (1.9) | 0.24 (0.04, 1.03) | 0.16 | 8 (7.8) | 1.47 (0.55,3.51) | 0.41 |
| High TC | No | 920 | 51 (5.5) | Ref |  | 29 (3.2) | Ref |  |
|  | Yes | 186 | 14 (7.5) | 1.24 (0.62, 2.36) | 0.53 | 20 (10.8) | 3.45 (1.72,6.94) | <0.001 |
| High LDL | No | 894 | 53 (5.9) | Ref |  | 28 (3.1) | Ref |  |
|  | Yes | 212 | 12 (5.7) | 0.89 (0.43, 1.72) | 0.74 | 21 (9.9) | 3.35 (1.69,6.65) | <0.001 |
| Low HDL | No | 945 | 56 (5.9) | Ref |  | 40 (4.2) | Ref |  |
|  | Yes | 161 | 9 (5.6) | 0.70 (0.30, 1.47) | 0.38 | 9 (5.6) | 1.09 (0.43,2.49) | 0.85 |
| Dyslipidemia | No | 694 | 43 (6.2) | Ref |  | 19 (2.7) | Ref |  |
|  | Yes | 412 | 22 (5.3) | 0.69 (0.39, 1.20) | 0.20 | 30 (7.3) | 2.26 (1.19,4.37) | 0.01 |
| High Lp(a) | No | 867 | 52 (6.0) | Ref |  | 37 (4.3) | Ref |  |
|  | Yes | 239 | 13 (5.4) | 0.85 (0.43, 1.59) | 0.63 | 12 (5.0) | 0.88 (0.41,1.77) | 0.73 |
| High hsCRP | No | 671 | 43 (6.4) | Ref |  | 25 (3.7) | Ref |  |
|  | Yes | 435 | 22 (5.1) | 0.71 (0.40, 1.24) | 0.24 | 24 (5.5) | 1.29 (0.68,2.44) | 0.43 |
| Hyperuricemia | No | 964 | 58 (6.0) | Ref |  | 26 (2.7) | Ref |  |
|  | Yes | 142 | 7 (4.9) | 0.73 (0.28, 1.62) | 0.47 | 23 (16.2) | 5.56 (2.79,11.15) | <0.001 |

Logistic regression model was used for statistical analysis. For overall obesity and abdominal obesity, age, sex, ethnicity, living area, education, smoking, and physical activity were adjusted in the model. For the other categorized risk factors, BMI was additionally added into the model. TG, triglyceride; TC, total cholesterol; LDL, low-density lipoprotein cholesterol; HDL, high-density lipoprotein cholesterol; Lp(a), lipoprotein (a); PAD, peripheral artery disease; LVH, left ventricular hypertrophy; CKD, chronic kidney disease; TOD, target organ damage.

**Figure 1.**
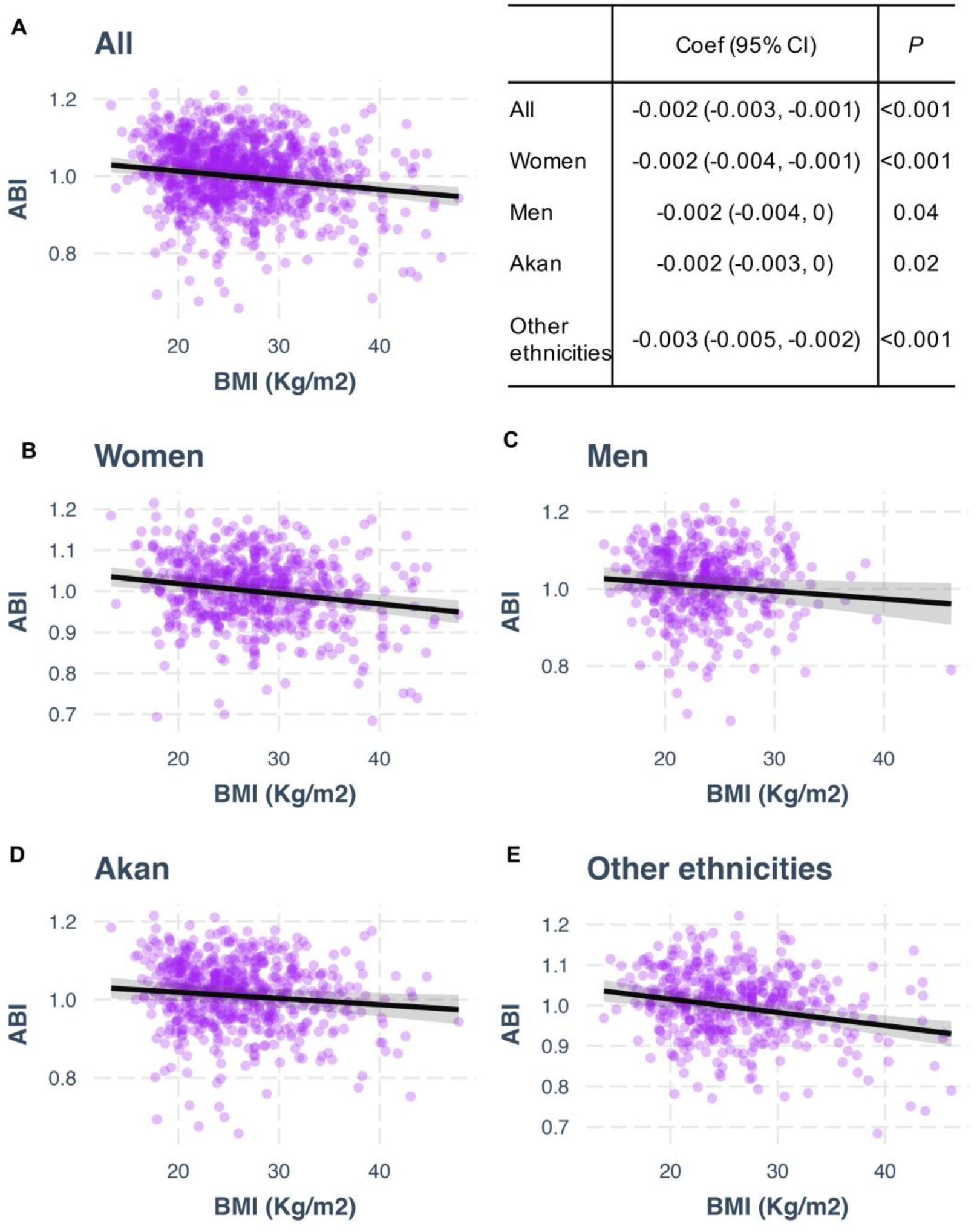
Association between body mass index (BMI) and ankle-brachial index (ABI) among adults participated in Ghana heart study, 2016–2017. Multivariate linear regression model was used for association analysis with adjustment for age, sex, ethnicity, living area, education, smoking, and physical activity.

Hypertension had the strongest association with carotid intimal thickening (OR 1.57, 95%CI 0.99–2.54), although the *P* value did not reach the statistically significant level. SBP and diastolic blood pressure (DBP) were significantly and positively associated with CIMT, after adjustment for potential confounding factors (Supplemental Table 1, Figure 2). Stratification analysis showed that DBP was significantly associated with CIMT in men but not in women, and the association between BP and CIMT existed only in Akan but not in other ethnicities (Figure 2).

**Figure 2.**
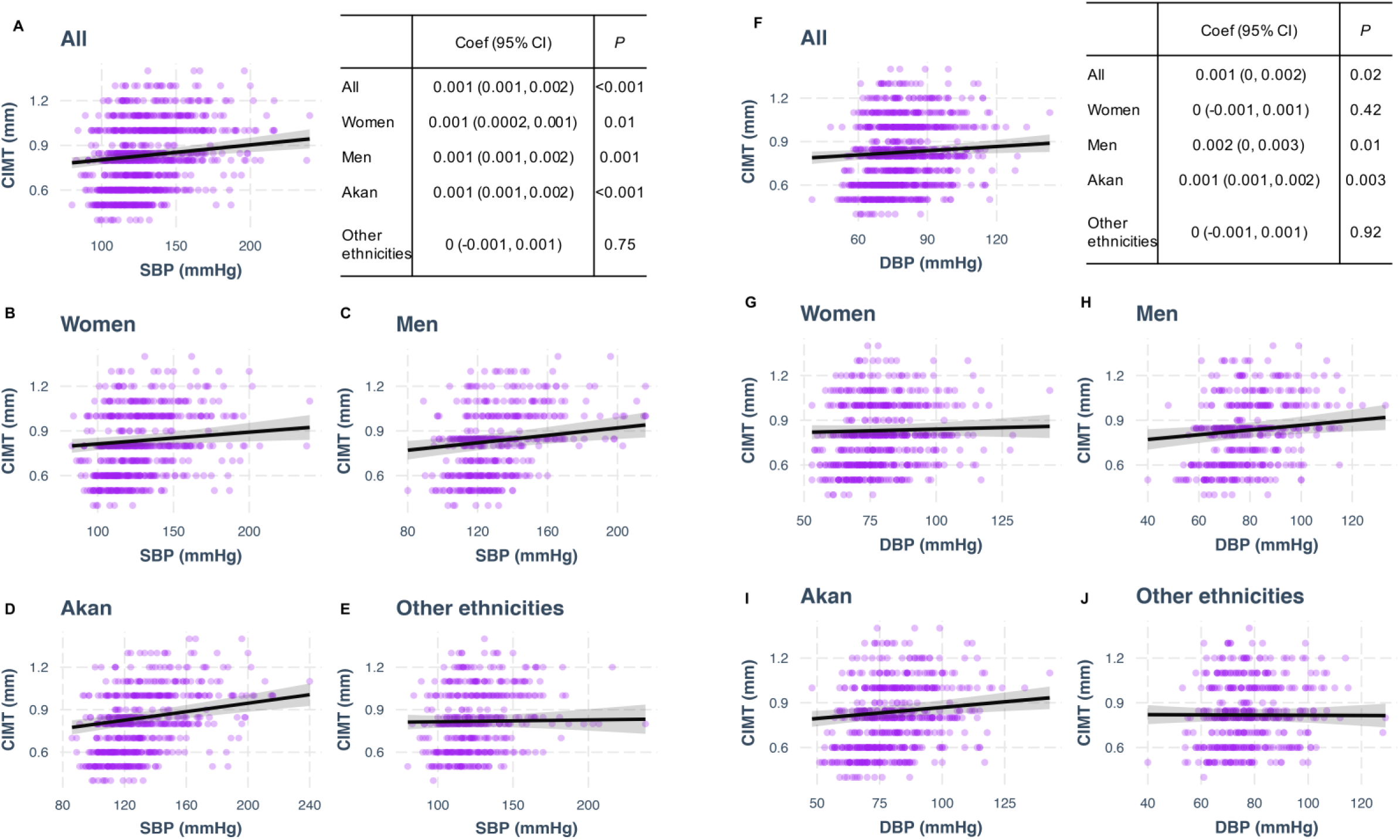
Association of systolic blood pressure (SBP) and diastolic blood pressure (DBP) with carotid intima–media thickness (CIMT) among adults participated in Ghana heart study, 2016–2017. Multivariate linear regression model was used for association analysis with adjustment for age, sex, ethnicity, living area, education, smoking, physical activity, and BMI.

Hypertension was strongly associated with LVH (OR 6.25, 95%CI 2.98–14.50) after adjustment for potential confounding factors (Table 2). Both SBP and DBP were positively and significantly associated with LVMI in the overall population after adjustment for potential confounding factors (*P* < 0.001). Stratification analysis showed that LVMI was associated with BP in all sex and ethnicities (Figure 3, Supplemental Table 1).

**Figure 3.**
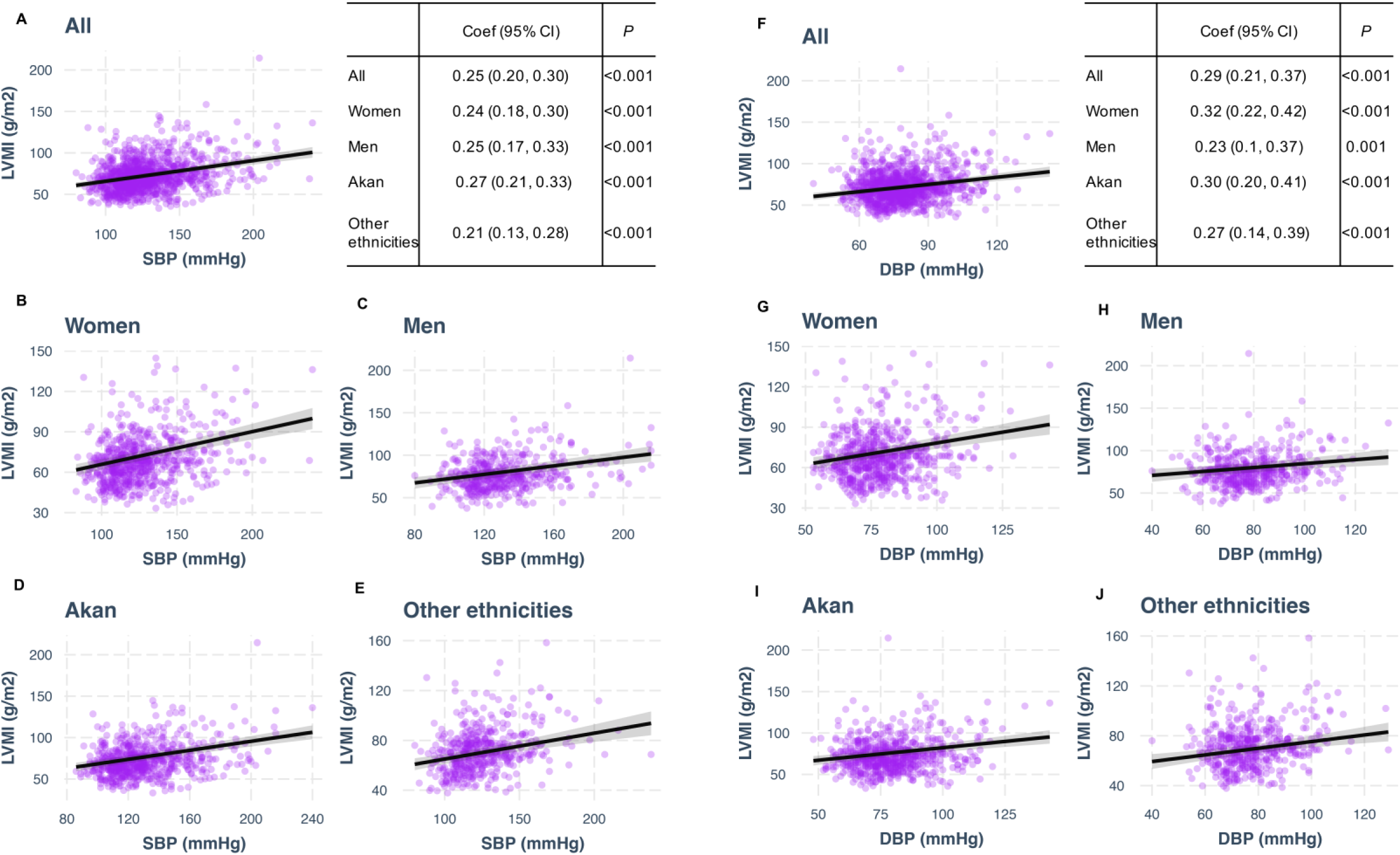
Association of systolic blood pressure (SBP) and diastolic blood pressure (DBP) with left ventricular mass index (LVMI) among adults participated in Ghana heart study, 2016–2017. Multivariate linear regression model was used for association analysis with adjustment for age, sex, ethnicity, living area, education, smoking, physical activity, and BMI.

Hyperuricemia had the strongest association with CKD (OR 5.56, 95%CI 2.79–11.15) after adjustment for potential confounding factors (Table 2). Uric acid was most strongly associated eGFR after adjustment for potential confounding factors. Stratification analysis showed significant associations of eGFR with uric acid in all sex and ethnicities (Figure 4, Supplemental Table 1).

**Figure 4.**
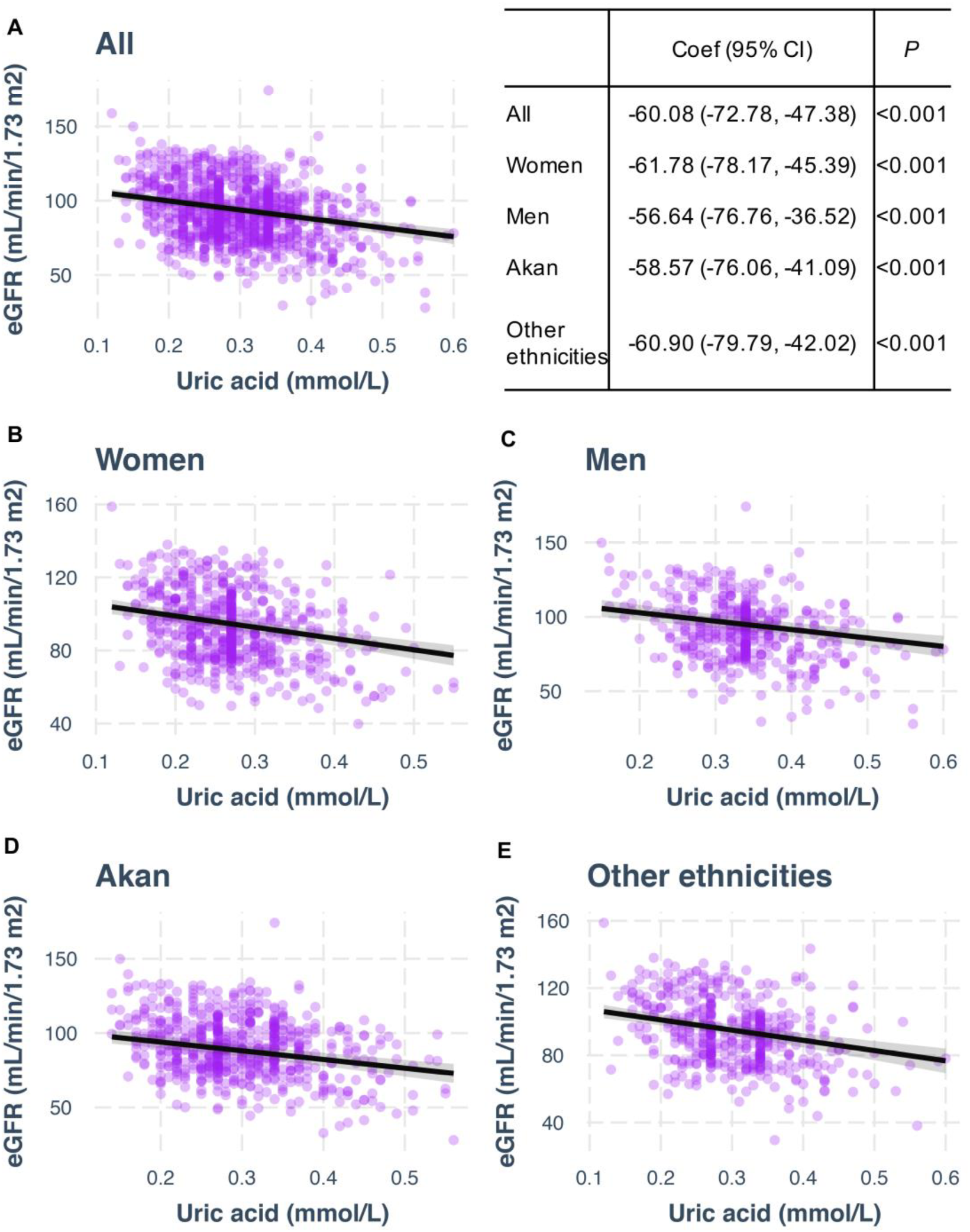
Association of uric acid with estimated glomerular filtration rate (eGFR) among adults participated in Ghana heart study, 2016–2017. Multivariate linear regression model was used for association analysis with adjustment for age, sex, ethnicity, living area, education, smoking, physical activity, and BMI.

## Discussion

Although CVD now claims more lives among adults in LMICs than in developed countries, very limited epidemiologic data are available in populations in LMICs. While the pathophysiological mechanisms underlying CVD are likely similar between populations in the developed countries and LMICs, the distribution of risk factors was most likely different in different regions across the world. This would determine what specific prevention and intervention strategy should be adopted for specific populations. In this study, we characterized the distribution of cardiometabolic risk factors and preclinical TOD in Ghana located in western SSA with the highest prevalence of CVD in the world. Our findings indicate that cardiometabolic risk factors previously identified in populations of the developed countries are now common among adults in Ghana. In particular, obesity, hypertension, and hyperuricemia appear to be the risk factors having the strongest association with specific preclinical TOD.

CVD has been recognized to arise from a chain of events since the 1990s. Termed the cardiovascular disease continuum (CVDC), the disease develops in association with a cluster of risk factors followed by asymptomatic preclinical TOD in organs such as blood vessels, the heart and kidneys, ultimately causing clinical CVD events ^7^. From the perspective of prevention, it is important to elucidate the association of the risk factors with preclinical TOD to facilitate interventions to disrupt the CVDC at the early stage. However, the contribution of the established cardiometabolic risk factors from studies in developed countries to CVD in LMICs has not been comprehensively investigated.

The prevalence of obesity, hypertension, diabetes, and dyslipidemia, all highly independent risk factors for CVD have been increasing globally. Our findings suggest that cardiometabolic risk factors are no longer rare in SSA. Specifically, we find that the obesity rate in Ghanaian adults is as high as 21%, and that there are clear differences between women (30%) and men (8%). The current prevalence of hypertension in our study is ~ 40% based on JNC7 or ~ 55% based on 2017 AHA guideline, which is comparable to that in the US or China^8 9^.Notably, the current study shows that type 2 diabetes (T2D) prevalence is now 11% among adults in Ghana, as compared to studies conducted from 1960s to 1980s showing T2D prevalence of < 1% in many regions of SSA^10^. This estimate of T2D prevalence in Ghanaian adults is comparable to the prevalence rate observed in populations in other regions of the world ^11^.Our findings clearly show that dyslipidemia is a common risk factor in Ghanaian adults, which is consistent with one recent meta-analysis demonstrating that dyslipidemia is not rare in Africa but has a high prevalence in the general adult population in Africa^12^.

Uric acid is the end product of purine metabolism in human body. Although the causal relationship between uric acid and CVD remains unclear, many epidemiological studies suggest the existence of a significant association between high uric acid and increased risk of CVD^13^ as well as CKD^14^. In our study, hyperuricemia was defined according to the criteria adopted by the NHANES. Our data indicate that ~13% adults are hyperuricemic in Ghana. As a comparison, hyperuricemia prevalence rates were 20% among adults in the US in 2015–2016^15^.

Inflammation has been thought to play key roles in the pathophysiology of CVD. We also measured plasma hsCRP as a marker of inflammation in the current study. Despite publication of guidelines on the use of hsCRP in the primary prevention of CVD ^16 17^, the optimal clinical use of hsCRP remains controversial ^18^. Previous studies in African populations have investigated the relationship between inflammation and infectious diseases ^19^, however, the association between hsCRP and CVD has not been well studied in SSA. A study in Burkina Faso suggests that intervention targeting on hsCRP could be effective in preventing CVD events in SSA ^20^. Our study finds that the prevalence of high hsCRP in Ghana is very similar to that of Burkina Faso, a geographical neighbor.

We also found clear sex and ethnicity discrepancy in CVD risk factors prevalence in Ghana. Women had higher BMI, waist, TC, LDL, HDL, and hsCRP, but lower SBP and uric acid than men. Akan had significant higher diabetes, hypertension, and lipids than other ethnicities. Further investigations are needed to clarify the reasons causing such discrepancy and whose effects on CVD development in Ghana.

We further investigated the roles of CVD risk factors in the development of preclinical conditions of vascular, cardiac, and renal TOD. PAD and carotid thickening were chosen as the proxy measurements for vascular TOD. A recent systematic review shows that the prevalence of PAD in LMIC is 3% at age 45–49 years and 15% at age 85–89 years, which is lower than in high-income countries with smoking, diabetes, hypertension, and high TC are identified as the risk factors for PAD ^21^. Our study showed that the prevalence of PAD was ~9% in the Ghanaian adult population with mean age of 47 years. We identified obesity and high TC as the most important risk factors for PAD in Ghana. Also, we found that the prevalence of carotid thickening was ~ 15% in this population and hypertension was considered to be the key risk factor, consistent with the previous report ^22^.

The cardiac TOD was evaluated by LVH in our samples. It has been reported that blacks are more likely to develop LVH than whites in the United States^23^. The studies on black populations living in Africa show that the overall prevalence of LVH in Gambia was 41%^24^, 62% in Cameroon^25^, and 41% in Angola^26^.Our study indicated that the prevalence of LVH in Ghana (~6%) is significantly lower than that in other African countries. Further studies with comprehensive comparison of sociodemographic and cultural factors in Ghana versus the other African countries are warranted. Consistent with the other studies^27^, we found that hypertension has the strongest association with LVH in Ghanaian population.

While the prevalence of CKD was low in our population (4.4%), high TC, high LDL, and hyperuricemia were significantly associated with CKD. The prevalence of CKD was 13% in the US according to the NHANES 1999–2004 data ^28^ and 11% in China in 2012 showed in a national survey^29^. The China national survey reported significant association of hypertension, diabetes, and hyperuricemia with CKD. Although hyperuricemia has a strong association with the risk of CKD (OR=5.6 in our study; OR=9.3 in the China survey), a recent mendelian randomization study concluded no causal effects of serum urate level on the risk of CKD ^30^. Thus. the strong association of hyperuricemia with risk of CKD observed in multiple populations need to be further investigated in future work.

While this is a cross-sectional study from which direct causal claim cannot be made, we wish to note that all CVD risk factors identified in prospective cohorts of men and women in the developed countries were confirmed. Importantly, for the first time, we were able to directly estimate the prevalence of preclinical target organ damage along with associated risk factors in Ghana, setting the stage for further prospective or intervention studies in this population.

In summary, in this first national survey of cardiometabolic health status among adults in Ghana, known CVD risk factors were found to be highly prevalent, at a par with those reported in the high-income countries. Specifically, obesity, hypertension, and hyperuricemia were important CVD risk factors associated with preclinical TODs among adults in Ghana which should be the focus for interventions to improve both individual- and population-cardiometabolic health in Ghana.

## Data Availability

The data can be available by contacting the corresponding authors.

## Perspectives

### Competency in systems-based practice

While the pathophysiological mechanisms underlying CVD are likely similar between populations in the developed countries and LMICs, the distribution of risk factors was most likely different in different regions across the world. Our study would be helpful for determining what specific prevention and intervention strategy should be adopted for specific populations in SSA.

### Translational outlook

More work is needed to examine if the prevention strategies directed at these risk factors can reduce the CVD burden in Ghana at both the individual and population level.

## References

1. Miranda JJ, Barrientos-Gutierrez T, Corvalan C, et al. Understanding the rise of cardiometabolic diseases in low- and middle-income countries. Nat Med 2019;25(11):1667–79.

2. Roth GA, Johnson C, Abajobir A, et al. Global, Regional, and National Burden of Cardiovascular Diseases for 10 Causes, 1990 to 2015. J Am Coll Cardiol 2017;70(1):1–25.

3. Holmes MD, Dalal S, Volmink J, et al. Non-communicable diseases in sub-Saharan Africa: the case for cohort studies. PLoS Med 2010;7(5):e1000244.

4. Dzau VJ, Antman EM, Black HR, et al. The cardiovascular disease continuum validated: clinical evidence of improved patient outcomes: part I: Pathophysiology and clinical trial evidence (risk factors through stable coronary artery disease). Circulation 2006;114(25):2850–70.

5. Cannon CP. Cardiovascular disease and modifiable cardiometabolic risk factors. Clin Cornerstone 2007;8(3):11–28.

6. Buuren Sv, Groothuis-Oudshoorn K. mice: Multivariate imputation by chained equations in R. Journal of statistical software 2010: 1–68.

7. Dzau V, Braunwald E. Resolved and unresolved issues in the prevention and treatment of coronary artery disease: a workshop consensus statement. Am Heart J 1991;121(4 Pt 1):1244–63.

8. Fryar CD, Ostchega Y, Hales CM, et al. Hypertension Prevalence and Control Among Adults: United States, 2015–2016. NCHS Data Brief 2017(289):1–8.

9. Wang Z, Chen Z, Zhang L, et al. Status of Hypertension in China: Results From the China Hypertension Survey, 2012–2015. Circulation 2018;137(22):2344–56.

10. Mbanya JC, Motala AA, Sobngwi E, et al. Diabetes in sub-Saharan Africa. Lancet 2010;375(9733):2254–66.

11. Wang L, Gao P, Zhang M, et al. Prevalence and Ethnic Pattern of Diabetes and Prediabetes in China in 2013. JAMA 2017;317(24):2515–23.

12. Noubiap JJ, Bigna JJ, Nansseu JR, et al. Prevalence of dyslipidaemia among adults in Africa: a systematic review and meta-analysis. Lancet Glob Health 2018;6(9):e998-e1007.

13. Sarfo FS, Akassi J, Antwi NK, et al. Highly Prevalent Hyperuricaemia is Associated with Adverse Clinical Outcomes Among Ghanaian Stroke Patients: An Observational Prospective Study. Ghana Med J 2015;49(3):165–72.

14. Kumagai T, Ota T, Tamura Y, et al. Time to target uric acid to retard CKD progression. Clin Exp Nephrol 2017;21(2):182–92.

15. Chen-Xu M, Yokose C, Rai SK, et al. Contemporary Prevalence of Gout and Hyperuricemia in the United States and Decadal Trends: The National Health and Nutrition Examination Survey, 2007–2016. Arthritis Rheumatol 2019;71(6):991–99.

16. Genest J, McPherson R, Frohlich J, et al. 2009 Canadian Cardiovascular Society/Canadian guidelines for the diagnosis and treatment of dyslipidemia and prevention of cardiovascular disease in the adult - 2009 recommendations. Can J Cardiol 2009;25(10):567–79.

17. Greenland P, Alpert JS, Beller GA, et al. 2010 ACCF/AHA guideline for assessment of cardiovascular risk in asymptomatic adults: a report of the American College of Cardiology Foundation/American Heart Association Task Force on Practice Guidelines. J Am Coll Cardiol 2010;56(25):e50–103.

18. Yousuf O, Mohanty BD, Martin SS, et al. High-sensitivity C-reactive protein and cardiovascular disease: a resolute belief or an elusive link? J Am Coll Cardiol 2013;62(5):397–408.

19. Imrie H, Fowkes FJ, Michon P, et al. Low prevalence of an acute phase response in asymptomatic children from a malaria-endemic area of Papua New Guinea. Am J Trop Med Hyg 2007;76(2):280–4.

20. Zeba AN, Delisle HF, Rossier C, et al. Association of high-sensitivity C-reactive protein with cardiometabolic risk factors and micronutrient deficiencies in adults of Ouagadougou, Burkina Faso. Br J Nutr 2013;109(7):1266–75.

21. Fowkes FG, Rudan D, Rudan I, et al. Comparison of global estimates of prevalence and risk factors for peripheral artery disease in 2000 and 2010: a systematic review and analysis. Lancet 2013;382(9901):1329–40.

22. Roman MJ, Pickering TG, Pini R, et al. Prevalence and determinants of cardiac and vascular hypertrophy in hypertension. Hypertension 1995;26(2):369–73.

23. Drazner MH, Dries DL, Peshock RM, et al. Left ventricular hypertrophy is more prevalent in blacks than whites in the general population: the Dallas Heart Study. Hypertension 2005;46(1):124–9.

24. Nkum BC, Micah FB, Ankrah TC, et al. Left ventricular hypertrophy and insulin resistance in adults from an urban community in The Gambia: cross-sectional study. PLoS One 2014;9(4):e93606.

25. Jingi AM, Noubiap JJ, Kamdem P, et al. Determinants and improvement of electrocardiographic diagnosis of left ventricular hypertrophy in a black African population. PLoS One 2014;9(5):e96783.

26. Baldo MP, Goncalves MA, Capingana DP, et al. Prevalence and Clinical Correlates of Left Ventricular Hypertrophy in Black Africans. High Blood Press Cardiovasc Prev 2018;25(3):283–89.

27. Heckbert SR, Post W, Pearson GD, et al. Traditional cardiovascular risk factors in relation to left ventricular mass, volume, and systolic function by cardiac magnetic resonance imaging: the Multiethnic Study of Atherosclerosis. J Am Coll Cardiol 2006;48(11):2285–92.

28. Coresh J, Selvin E, Stevens LA, et al. Prevalence of chronic kidney disease in the United States. JAMA 2007;298(17):2038–47.

29. Zhang L, Wang F, Wang L, et al. Prevalence of chronic kidney disease in China: a cross-sectional survey. Lancet 2012;379(9818):815–22.

30. Jordan DM, Choi HK, Verbanck M, et al. No causal effects of serum urate levels on the risk of chronic kidney disease: A Mendelian randomization study. PLoS Med 2019;16(1):e1002725.

